# The dangers of data double dipping in assessing the classification accuracies of blood biomarkers in Alzheimer’s disease and related disorder research

**DOI:** 10.64898/2026.05.22.26353848

**Authors:** Tianshu Liu, Xuemei Zeng, Beth E. Snitz, Thomas K. Karikari, Rebecca A. Deek

**Author notes:** **Corresponding author:** Rebecca A. Deek, Department of Biostatistics and Health Data Science, School of Public Health, University of Pittsburgh, 130 De Soto Street, Pittsburgh, PA, 15261.

## Abstract

Blood biomarker models are increasingly used in Alzheimer’s disease and related dementia translational research, but predictive performance can be inflated when the same dataset is used for both model development and evaluation. We assess the effect of data double dipping using simulations and NULISA proteomic data from the MYHAT-NI community-based cohort to predict brain amyloid-beta neuroimaging status. In both settings, training AUC increased as more biomarkers were added, while testing AUC peaked earlier and then declined. These findings show that data double dipping can inflate model performance and highlight the need for external validation or internal validation with data partitioning.

## Introduction

In biomedical research, predictive models that incorporate either a single biomarker or multiple biomarkers with demographic variables are frequently used to predict a phenotype of interest, including disease status or progression [1, 2]. In neuroscience, it is of particular interest to link molecular biomarkers to cognitive status, Alzheimer’s disease (AD), or related dementias (ADRD) status, as measured by neuropathology, positron emission tomography (PET), or magnetic resonance imaging (MRI) [3, 4, 5, 6]. Given the growing popularity of blood biomarkers in ADRD research, many studies have been conducted to link these biomarkers with outcomes, such as amyloid-beta PET (Aβ-PET) status [7]. Such studies typically fit a classification model, such as a logistic regression model, to the data and evaluate its performance with prediction metrics such as area under the curve (AUC), sensitivity, specificity, accuracy, and F1 score [8]. An analytical concern is whether the same dataset can be used to “build” the model (i.e., estimate model parameters) and to validate the model in terms of predictive performance. This practice is sometimes referred to as “data double dipping”, as we have used the same data twice in building and validating the model [9, 10]. This can result in overly optimistic performance metrics since the model already “knows” the true class label for the data it was fit to during model training and stands against the objective of assessing classification performance on new, unseen data that does not contain any outcome label information, as this is how the model will be implemented in clinical practice.

This issue stands to grow as the number of biomarkers collected on individuals increases, as is the case with the rise of multiplex assay technologies. Due to the large number of biomarkers collected, and believed to be potentially associated with disease status, as well as the uncertainty about which are genuinely relevant, it is common for researchers to fit models that include all available biomarkers as predictors. Including more predictors results in explaining more variability of the outcome, as we are adding new knowledge to the model with each predictor. However, one should be cautious of the optimistically increasing performance metrics, especially when double dipping, as they can be inflated due to overfitting. Overfitting occurs when a model begins to learn the noise in training data used to build the model, which is unrelated to the outcome, and uses it to gain better predictive performance. When this occurs, we cannot identify whether the model is overfit until we apply it to new, unseen data and assess the prediction performance in this independent dataset. This is because the noise structure differs across samples. One way to identify overfitting is if there is a substantial drop in the prediction metrics between the two data sets. This is an indication that the model was leveraging some of the training data’s noise, thereby limiting its generalizability to new data.

Ideally, new data from the same target population would be collected for model validation. Realistically, due to constraints of real-world studies, this may not be feasible. In this case, data splitting, where the data is split into a training set for model building and a separate validation/testing set for evaluation of predictive performance, can be implemented prior to the model fitting step. Alternatively, we can consider cross-validation techniques, such as k-fold cross-validation, which repeatedly partition the data into training and testing subsets and average performance across partitions.

### Illustrative examples using simulated and real-world data

We illustrate the dangers of not utilizing separate training and testing data for predictive modeling in ADRD studies through simulation studies and real-world data. We use simulation studies in which we intentionally include additional predictors that are uncorrelated with a binary outcome, such as disease status, and therefore should not improve discrimination between “cases” (i.e., those with disease) and “controls” (i.e., those without disease). By incrementally adding these uninformative predictors, we create increasing degrees of overfitting. Given that the true data generating mechanism is known in simulation, the best-performing model should include all truly associated predictors and exclude unassociated predictors.

We focus on data splitting, as it is often the case that an external validation dataset from the same study population is not available. We use a 70/30 data partition, with 70% of the generated data used for model training and 30% used as a validation or testing data set that is fully left out of the model building procedure. We fit a sequence of logistic regression models, adding one additional predictor at a time, until all candidate predictors are included. For each model, we compute the AUC on the training data set and on the testing data set. The training AUC reflects performance evaluated on data that were used for model development and is therefore subject to double dipping. The testing set AUC reflects performance on unseen data, reflecting its generalizable performance to real-world clinical settings, and can reveal when a model overfits.

In each simulation replicate, we first generate data with sample size of *n* = 200 and a total of *p* mutually independent continuous predictors drawn from a multivariate normal distribution with mean zero and identity covariance. Only *k* (*k* < *p*) predictors are truly predictive of the binary outcome, ensuring that the remaining *p* – *k* predictors are not contributing to the binary outcome. We then split the data into a training set (*n* = 140) and a testing set (*n* = 60). During model training and building, we fit a sequence of logistic regression models starting with one truly contributing predictor and adding one additional predictor at each step, until all predictors are included. As the true value of *k* is known in simulations, we expect models with fewer than *k* predictors to underfit, the model with *k* predictors to be well-specified, and models with more than *k* predictors to become increasingly overfit as additional uninformative predictors are added. We repeat this process 500 times and summarize the mean training and testing AUC as a function of the number of predictors included in the model.

We consider two scenarios. In scenario (I), three of the fifteen predictors are truly associated with the outcome, and have true regression coefficients ***β*** = (1, 1.3, 0.7). In scenario (II), five of the forty predictors are truly associated with the outcome, with true regression coefficients ***β*** = (1, 1.3, 0.7, 1.2, 0.8).

In scenario (I), where only three predictors were truly related to the outcome, both training and testing AUCs increased as these three predictors were added into the logistic regression model first, reflecting improved model specification by learning new outcome-related information from every newly added predictor. After adding the third predictor to the model, which was expected to be the optimal fit, the mean training AUC was 0.847 and the mean testing AUC reached its elbow point near 0.837 (Figure 1). When more unrelated predictors were added to the model, the training AUC continued optimistically increasing gradually because the model kept learning noise patterns from the newly added predictors, while the testing AUC decreased steadily, indicating the model started overfitting and learning outcome-uninformative noise from the training data. In this setting, if only training AUC was assessed we would conclude overly optimistic model performance.

**Figure 1.**
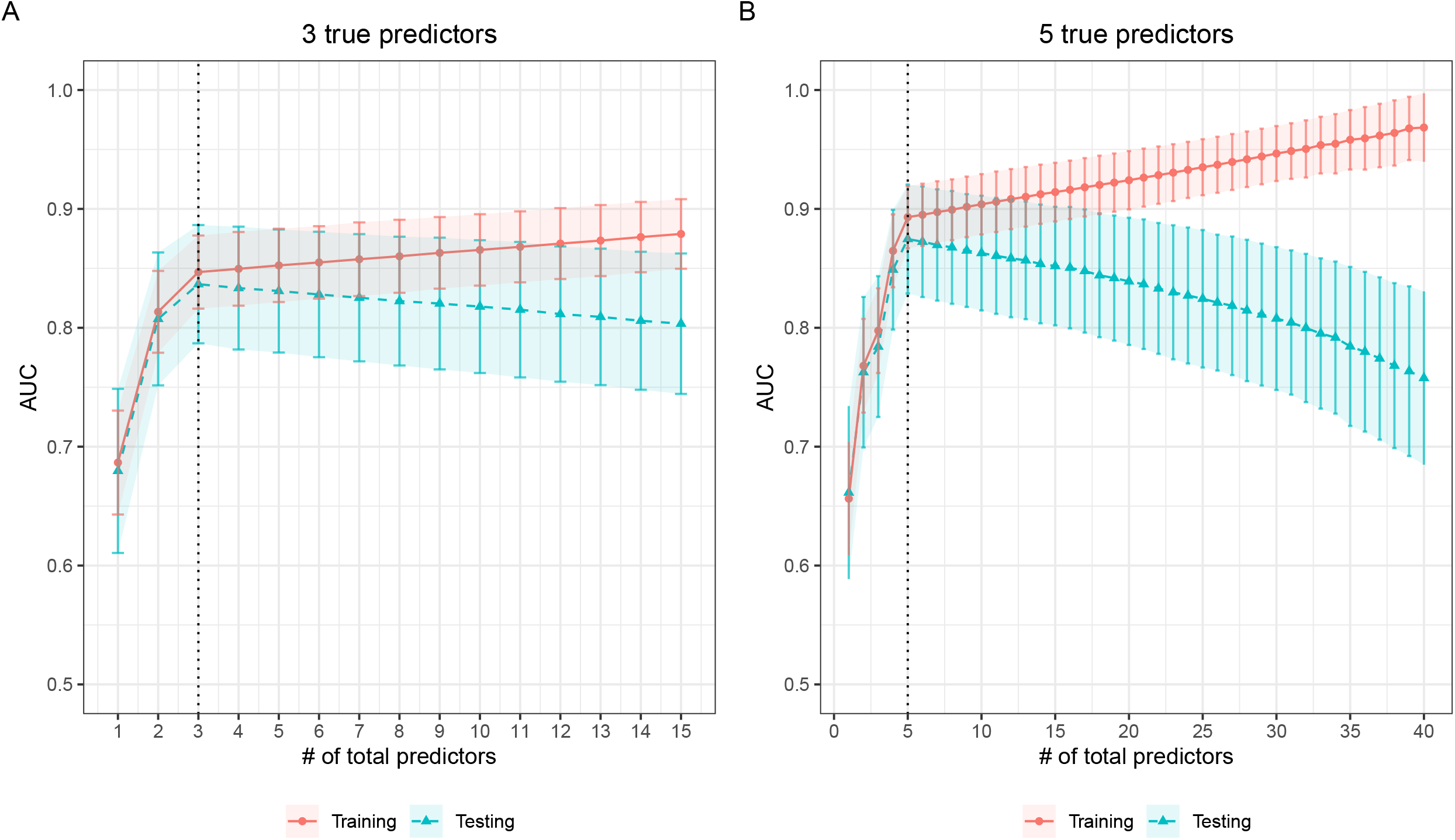
Training and testing AUCs in simulations. **(A)** Training and testing AUCs in scenario (I), where three of fifteen predictors are truly associated with the outcome. (**B**) Training and testing AUCs in scenario (II), where five of forty predictors are truly associated with the outcome. Points denote the mean AUC across simulation replicates and error bars indicate variability across replicates. Training AUC (red) continues to increase as uninformative predictors are added, whereas testing AUC (blue) peaks near the correctly specified model and then declines.

In scenario (II), which included five total true predictors, the AUC trends were similar. The main difference was that the training AUC and testing AUC were higher compared to the scenario (I) at the expected optimal fit, and the training AUC approached its maximum value of 1, as more unrelated predictors were included (Figure 1).

To illustrate these issues in real data, we analyzed data from the Monongahela– Youghiogheny Healthy Aging Team Neuroimaging (MYHAT-NI) cohort, a subset of the ongoing MYHAT prospective aging cohort. The MYHAT study recruited adults aged 65 or older from the Rust Belt region beginning in 2006, with another wave in 2016–2019 [11]. The MYHAT-NI cohort enrolled a subset of participants from the population-based MYHAT study for two-year longitudinal follow-up of Aβ, tau, and neurodegeneration using PET and structural MRI between 2016 and 2019 [12]. In MYHAT-NI, Aβ positivity was defined as a mean standard uptake value ratio (SUVR) > 1.346, and participants had CDR sum-of-boxes < 1.0 at enrollment. Sociodemographic variables were collected at the baseline neuroimaging visit. Blood sampling, neurophysiological assessment, and neuroimaging were performed at both baseline and two-year follow-up. For the present analysis, we used a multiplex NULISA assay dataset comprising 116 continuous blood-based biomarkers and a binary Aβ-PET status indicator for 111 participants at their baseline visit, where 28 (25.23%) are Aβ-PET positive. As most of the biomarkers were correlated to the binary outcome, it was plausible to include them in a multivariate model to predict the binary Aβ-PET status. To assess whether adding more biomarkers improves logistic regression prediction performance, we split the data into training and testing sets, selected the 25 biomarkers with the largest absolute point-biserial correlation with the binary outcome, and then added these biomarkers incrementally to models fitted on the training set. We computed the AUC for both training set and testing set at each step. Consistent with the findings in our simulation experiment above, training AUC increased almost monotonically as predictors were added, while testing AUC peaked early and then declined. The testing AUC had its global peak when the model included four predictors: p-tau217, gfap151, p-tau231, and timp3, indicating an overfitting issue of the model after this point (Figure 2). This result emphasizes the importance of data partitioning in practice: the training set performance is not sufficient to detect model overfitting, as the prediction performance can always be overly optimistic due to data double dipping. In contrast, the testing set performance can tell promptly when the model starts to overfit and is more representative of the prediction model’s generalizable performance in real-world data settings.

**Figure 2.**
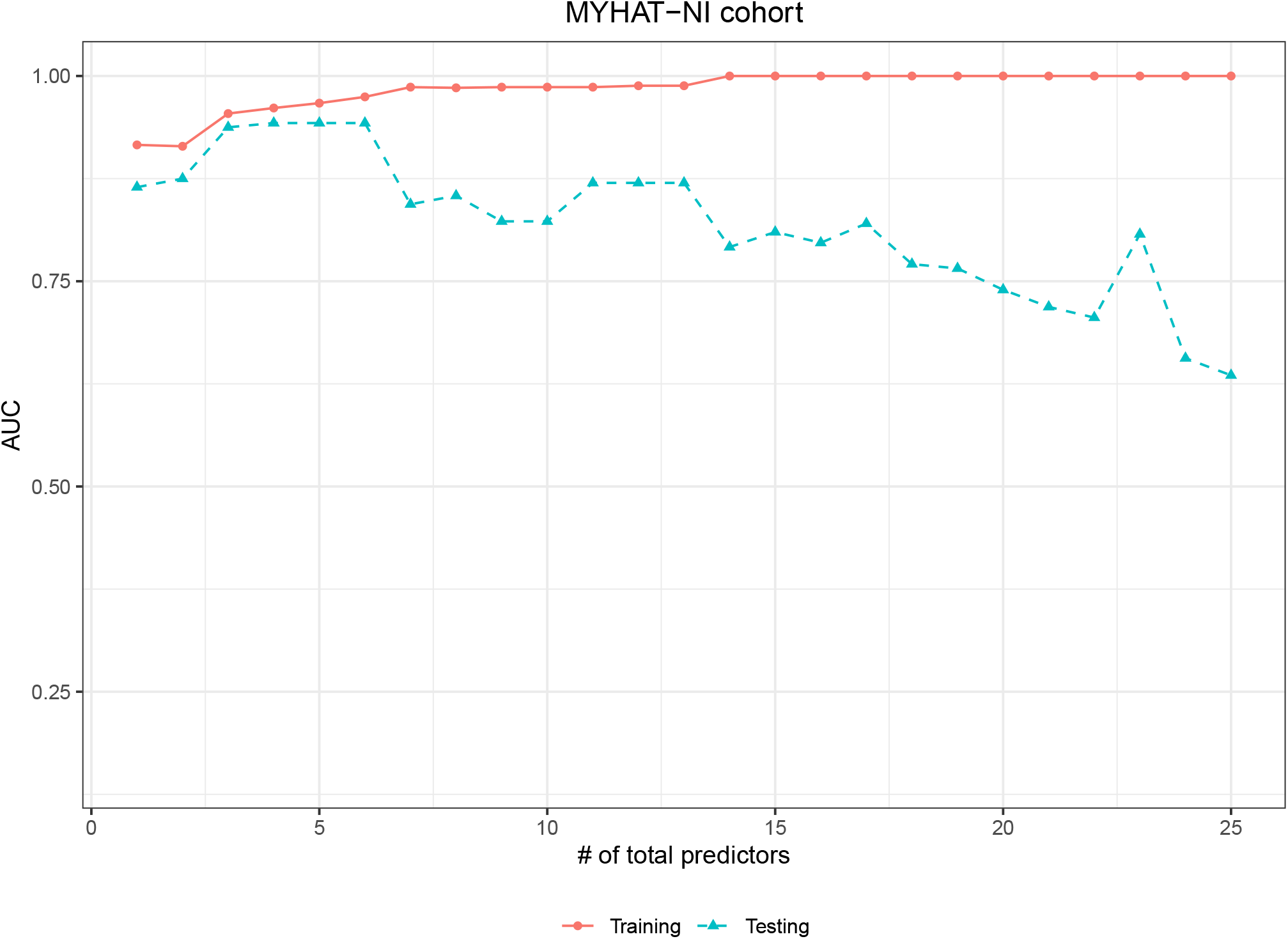
Training and testing AUCs for predicting Aβ-PET status in the MYHAT-NI cohort using multiplex NULISA proteomics for an increasing number of biomarker predictors. The x-axis represents models with an increasing number of predictors. The y-axis represents the model AUC. Training AUC (red) approaches its maximum value of 1 with additional predictors, while testing AUC (blue) peaks early and subsequently declines.

## Discussion

In this article, we demonstrate that assessment of predictive performance of blood biomarkers in ADRD studies must be evaluated on new, unseen data in clinical practice. Only when evaluated on an external dataset, unseen during model building, can we promptly assess generalizability and detect overfitting. Using simulations, we show that training AUC increases with model complexity, even when additional predictors are uninformative. In contrast, testing AUC reflects the performance on new data and shows a clear drop once uninformative predictors are added. The growing gap between the two AUCs is because the model can learn outcome-uninformative information from training data, including noise patterns, resulting in inflated performance when evaluated on the same data. Since noise patterns are random, and not replicable across independent dataset, the performance declines when evaluated on new data.

When using blood biomarker data from a real-world ADRD study, the truly outcome-associated predictors are unknown, but it is important to promptly detect overfitting early to preserve the accuracy and generalizability of prediction. This is particularly relevant in AD biomarker research, where high-dimensional biomarker panels and flexible modeling pipelines are increasingly being investigated and can yield erroneous performance estimates if model selection and evaluation are not separated. Thus, investigators should always avoid evaluating performance on the same data used for model development. Instead, an external validation set should be used when feasible or otherwise partition the available dataset into separate training and testing sets. Only prediction performance metrics on the separate testing set should be reported when discussing model generalizability. Moreover, carefully assessment of proper model validation techniques during the review process, by peer reviewers and editors alike, will support advances in ADRD biomarker research by ensuring more valid, generalizable, and replicable prediction metrics are reported.

## Acknowledgments

We thank the MYHAT-NI study participants, as well as their families and caregivers.

## Author Contributions

TL and RAD performed the data analysis. All authors contributed to writing the manuscript.

## Statements and Declarations

### Ethical considerations

All participants provided written consent, and the University of Pittsburgh Institutional Review Board approved the study.

### Consent to participate

All participants provided written consent, and the University of Pittsburgh Institutional Review Board approved the study.

### Consent for publication

All participants provided written consent, and the University of Pittsburgh Institutional Review Board approved the study.

### Data availability

The data to support the findings of this study are available upon reasonable request.

